# Predicting the severity of disease progression in COVID-19 at the individual and population level: A mathematical model

**DOI:** 10.1101/2021.04.01.21254804

**Authors:** Narendra Chirmule, Pradip Nair, Bela Desai, Ravindra Khare, Vivek Nerurkar, Amitabh Gaur

**Affiliations:** SymphonyTech Biologics, Philadelphia, Pennsylvania, USA; Biocon, Bangalore, Karnataka, India; NanoCellect Biomedical, Inc., San Diego, California, USA; Department of Tropical Medicine, Medical Microbiology and Pharmacology, John A. Burns School of Medicine, University of Hawaii at Manoa, Honolulu, Hawaii, USA; Innovative Assay Solutions LLC, San Diego, California, USA

**Keywords:** innate, interferon, cytokine-storm, lymphopenia, neutralizing-antibodies, viral factors, modeling, prediction

## Abstract

The impact of COVID-19 disease on health and economy has been global, and the magnitude of devastation is unparalleled in modern history. Any potential course of action to manage this complex disease requires the systematic and efficient analysis of data that can delineate the underlying pathogenesis. We have developed a mathematical model of disease progression to predict the clinical outcome, utilizing a set of causal factors known to contribute to COVID-19 pathology such as age, comorbidities, and certain viral and immunological parameters. Viral load and selected indicators of a dysfunctional immune response, such as cytokines IL-6 and IFNα, which contribute to the cytokine storm and fever, parameters of inflammation d-dimer and ferritin, aberrations in lymphocyte number, lymphopenia, and neutralizing antibodies were included for the analysis. The model provides a framework to unravel the multi-factorial complexities of the immune response manifested in SARS-CoV-2 infected individuals. Further, this model can be valuable to predict clinical outcome at an individual level, and to develop strategies for allocating appropriate resources to mitigate severe cases at a population level.

## INTRODUCTION

The COVID-19 pandemic caused by infection with SARS-CoV-2 was officially announced in March 2020 by the CDC and WHO [1, 2]. As of this publication, more than 100 million infections and over 2.6 million deaths have been reported worldwide. Majority of the subjects have asymptomatic infections. The rate of fatality is disproportionately high in the elderly and patients with comorbidities such as diabetes, cardiac disease, and kidney disease [3, 4]. The consequences of the pandemic are fraught with potential loss of lives, social and economic distress, and the uncertainty of disease progression because of variable individual pathogenesis.

A unique and dysregulated immune response has been shown to be a hallmark of COVID-19 [5-9]. **Figure 1** schematically depicts the cascade of events that contribute to the progression of disease. Mathematical models have been utilized by several investigators to understand the mechanisms of disease pathogenesis, immune pathways involved and course of viral infections [10, 11]. In this article, we have proposed a predictive model that utilizes the levels of clinical and laboratory parameters to determine the severity of clinical outcomes ranging from asymptomatic to mild, moderate, severe, and critical disease states. The proposed model can be useful to predict clinical outcome at the individual-level and develop efficient and effective treatment strategies to manage public health challenges at the population-level.

**Figure 1.**
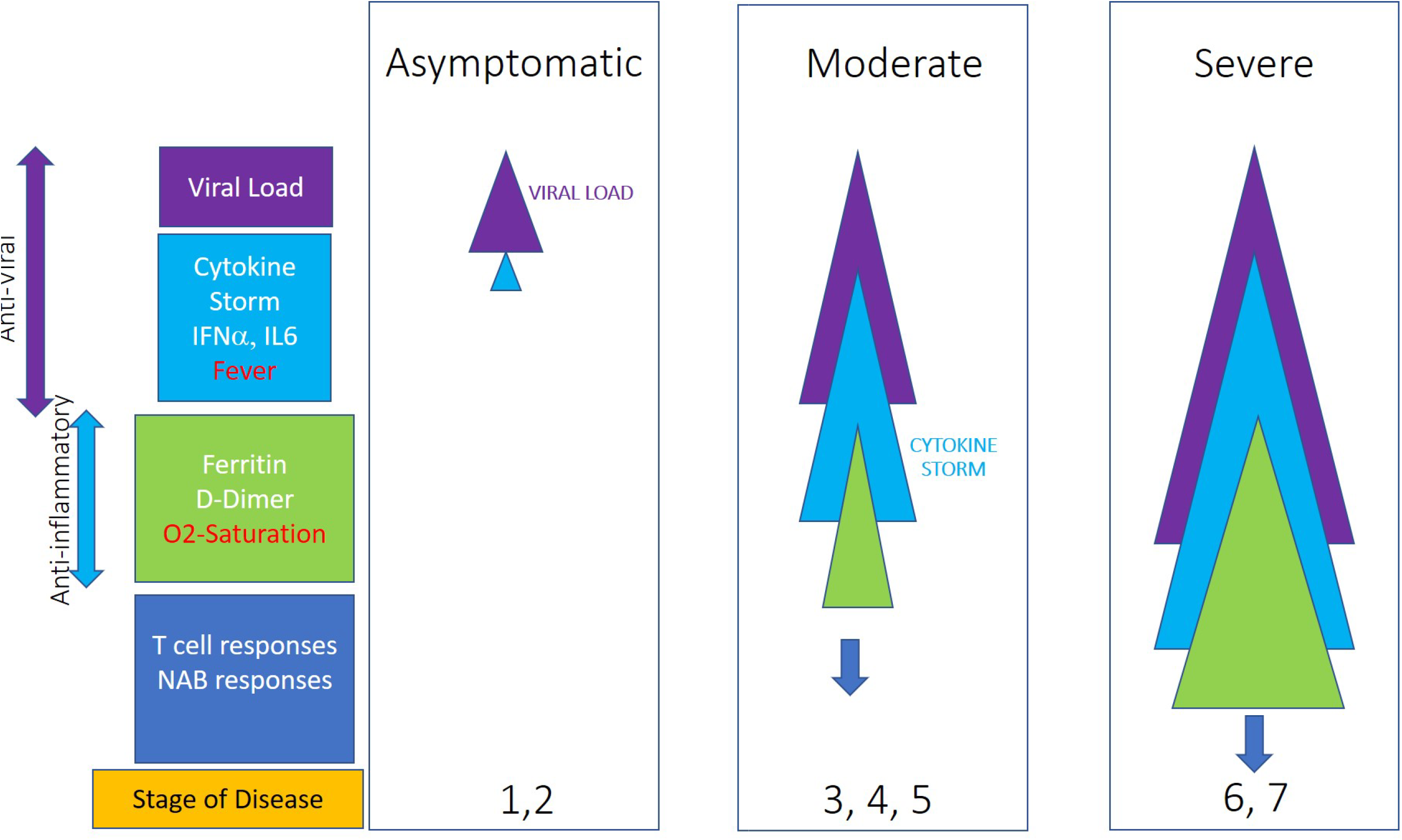
Schematic representation of the progression of disease. The width of the triangles denotes increase in levels of viral load (purple), cytokine storm (blue), and anti-inflammatory symptoms (green); blue arrows denote T and B cell responses.

The questions the model attempts to answer are: i) At an individual level, what is the probability of an individual infected with SARS-CoV-2, given the clinical signs and laboratory values on various days, likely to progress to severe disease, and ii) At a population level, what are the prioritized clinical and laboratory parameters that are most likely to contribute to progression to severe disease. We have used a multiple regression based model to predict severity of the outcome of COVID-19. To evaluate the combinatorics that are not observed in the sample, we have applied resampling methods based on Monte Carlo simulation.

## METHODS

### Development of a Simulated Dataset

A simulated data set of 45 individual subjects was created with 15 subjects assumed to be asymptomatic, 15 with moderate disease, and 15 with severe COVID-19 [12, 13]. The simulated values for the viral and immune parameters were generated using data from clinical reports published in the last year for each of the selected parameters. **Table 1** provides the ranges and the related references for the values for all parameters and **Figure 2** shows the box-and-whisker plots for the distribution of the values for each parameter.

**Table 1.** Ranges of values for the parameters used for developing the simulated dataset for the mathematical model. The range of comorbidities was assigned arbitrary nominal value between 1 to 4, with 1 being healthy, and 4 having multiple health-conditions (e.g., diabetes, cancer etc.). The age ranges in the model were 18-100 years.

| Parameter | Unit | Reference | COVID-19 Ranges | COVID-19 Ranges | COVID-19 Ranges |
| --- | --- | --- | --- | --- | --- |
|  |  |  | Asymptomatic | Moderate | Severe |
| **Viral Load** | Cycle Time | [20] | >28 | 20-15 | 22-16 |
| **IFNα** | pg/mL | [45, 46] | <10 | 10-100 | 10-2 |
| **Fever** | °F | [55, 56] | 97-98.6 | 98.6-100 | 100-104 |
| **D-Dimer** | $\mu\text{g/mL}$ | [31] | <0.1 | 0.15-0.62 | 0.5-9.3 |
| **Ferritin** | ng/L | [31] | 20-200 | 286-1,275 | 1,400-2,000 |
| **Oxygen Saturation** | % | [3, 56] | 95-100 | 85-94 | 60-84 |
| **IL-6** | ng/mL | [45, 46] | <1 | 19-76 | 19-430 |
| **Lymphocyte count** | $\times 10^6/\text{mL}$ | [27] | >785 | 588-785 | 169-415 |
| **NAB** | Titer | [57] | 1,000-45,000 | 200-20,000 | 500-60,000 |

**Figure 2.**
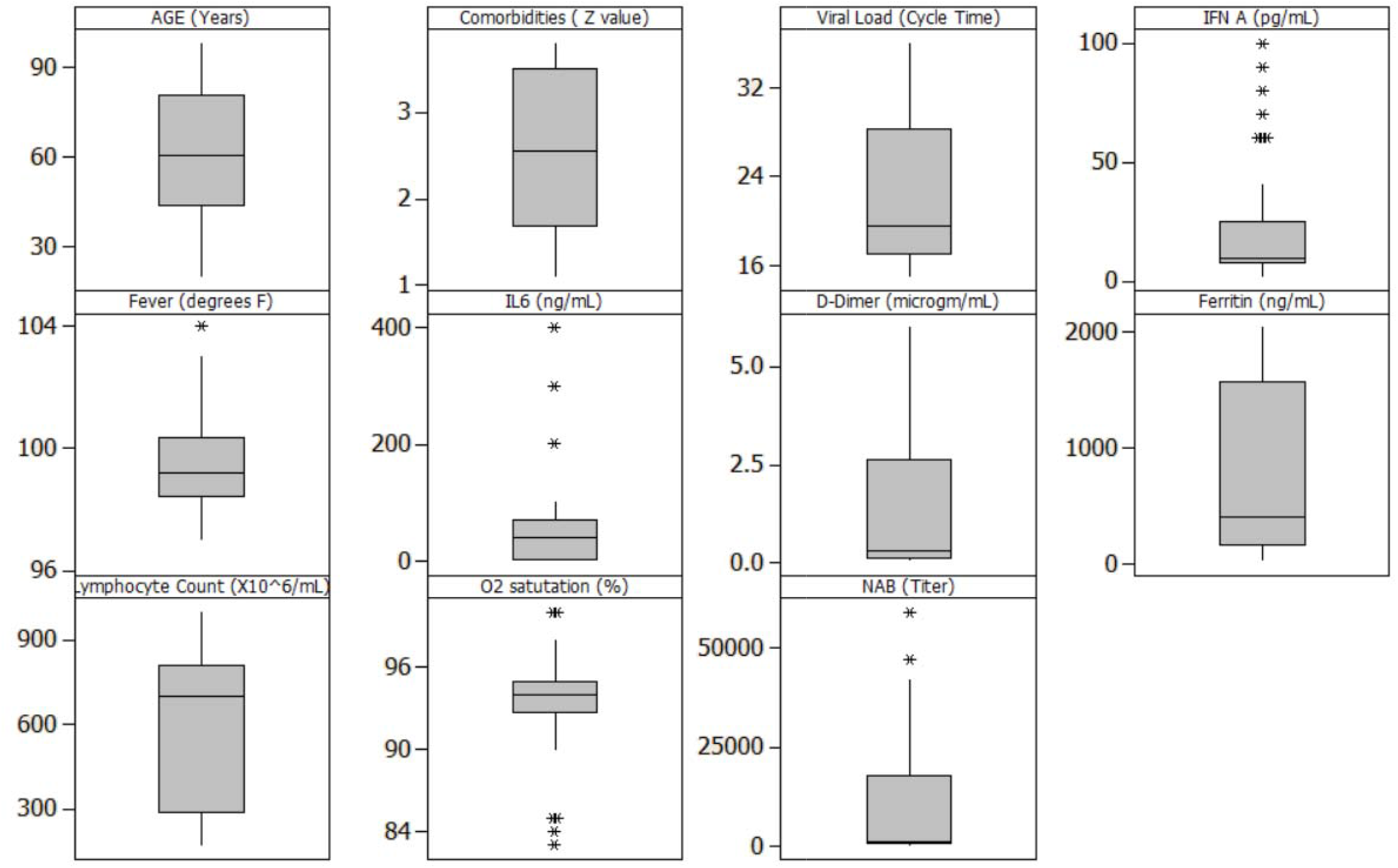
Box-and-whisker plots of the simulated data. The figures show the visual representation of the summary, which includes median (Q2/50^th^ percentile); first quartile (Q1/25^th^ percentile); third quartile (Q3/75h percentile); interquartile range in whiskers, maximum and outliers.

### Data Modeling

We have applied Multiple Linear Regression approach to the simulated data set for COVID-19 subjects generated and analyzed to understand the impact of each of the parameters on the outcome of disease severity. We chose a multiple regression model since both, the outcomes and predictors, were numeric. We used regression models to establish a predictive transfer function and evaluated significance of results. In this model, the relationship between independent variables (**x1, x2…x**_**n**_) with dependent variable (**y**) can be visualized by the equation, **y=f (x1, x2…x**_**n**_). This is the transfer function that is derived through analysis. The validity of the model was established using ‘Goodness of Fit’ and ANOVA. The statistical significance of the model was tested by evaluating residuals and F Ratio in one-way ANOVA, based on the criteria of p <0.05 and goodness-of-fit with Adj. RSq >90%. The assumption for this analysis was that each of the parameters was independent. However, in cases where factual patient datasets will be subjected to this type of analysis, there may be multi-co-linearity within the parameters that should be rationalized using dimensionality reduction methods [14, 15].

Since the model may not exhibit multiple combination of parameters in the limited dataset of 45 subjects, we have used resampling methods using Monte Carlo simulation to achieve a better density of combinations. The simulation was applied for resampling of the transfer function with 2000 runs, where a convergence was achieved after multiple runs. The simulation was performed in order to understand the impact of possible parameter combinations on clinical outcomes. Monte Carlo simulation uses random variates from selected range of values to model the impact of progression of events leading to outcomes.

### Data analysis using training and testing data sets

Model building involved partitioning the data set into ‘training’ and ‘testing’ sets. We apportioned 70% of the data to train the model and used the remaining 30% to test the model, using random selection algorithms. Following development of the model, we analyzed a set of test data to compare predicted versus observed results to validate the model. The regression model generated a prediction formula as follows:

**Outcome** = -36.898 - 0.020 AGE + 0.894 COMORBID - 0.048 Viral Load - 0.004 IFNα + 0.444 Fever - 0.003 IL6 + 0.271 D-Dimer + 0.000 Ferritin - 0.000 Lymphocyte Count - 0.037 O_2_ saturation - 2.57034e-006 NAB

The linear coefficients of the prediction equation determined the weights of each parameter to predict the clinical outcome.

### Estimation of the coefficients of input parameters

The modeling approach was based on utilizing clinical and laboratory parameters to fit the regression models. Since direct comparison of regression coefficients was not necessary, and interactions in factors were not considered on account of assumption of independence of factors, we chose to leave the factor-data in the original scales.

### Rationale for the parameters included in the analysis

The input parameters selected for this model, which requires cause (clinical and laboratory parameters) and effect (clinical outcome) relationships, were based on the data reported in recent scientific publications. **Figure 1** shows the schematic representation of the stage of disease progression and parameters associated with the increasing severity of diseases. The following parameters were chosen:

1. **Comorbidities:** Though the precise mechanism(s) of disease progression in patients with comorbidities has yet to be elucidated, pre-existing conditions such as diabetes, cancer, neurological, cardiac and lung and kidney disease have been reported to contribute towards severity of COVID-19 [16, 17]. The simulated data for comorbidity was generated using an arbitrary range of 1 to 4, where 1 represented a healthy individual and 4 represented an individual with a severe co-morbidity.
2. **Age**: A range of 18 to 100 years was utilized for generating the mock data set. The assumption used in generating the data was that disease progression was directly proportional to age [17]. Reports of certain rare pathogenic conditions in children, e.g., Kawasaki disease [18, 19], have not been considered in the current model. Reports indicate that majority of children infected with SARS-CoV-2 are asymptomatic [19].
3. **Viral load**: SARS-CoV-2 infects individuals through the nasopharyngeal pathway. This infection is the cause of all subsequent effects. Viral load is measured by reverse-transcriptase quantitative PCR (RT-qPCR), which detects viral RNA from nasopharyngeal swabs [20]. The test relies on multiple cycles of RNA amplification to produce detectable amount of RNA in the mixed nucleic acid sample, reflected in the Cycle-time (Ct) value, which is defined as the number of cycles necessary to detect the virus. A Ct value of less than 20 is considered a high viral load while a Ct value of 35 and higher indicates a lower level or near absence of viral infection [20]. Viral load in patients is dependent on various factors, including number of ACE2 and TMPRSS2 receptors [21, 22], comorbidities, cytokines, number of viral particles at infection, and the overall immune health status of the patients [23-26]. Viral loads have been demonstrated to have a direct correlation with severity of disease and mortality in COVID-19 [27, 28].
4. **Cytokine Storm**: High viral loads evoke defensive mechanisms that can induce inflammation leading to a dysregulated innate immune response that could result in a cytokine storm characterized by fever-inducing levels of cytokines such as IL6, IFNα, IL1β and CXCL-10 [27, 29-33]. CXCL-10, interestingly was also found to be indicative of severe outcomes in patients affected by the SARS CoV1 outbreak in 2002 [34]. Cytokine storm has been implicated in contributing to pulmonary immunopathology, leading to severe clinical disease and mortality. In this model, we have included levels of IFNα and IL6 obtained from the published data.
5. **Systemic Inflammation**: Laboratory based parameters indicating inflammation in the serum, such as D-Dimer and Ferritin, have been shown to lead to a reduction in blood oxygen saturation levels, reflecting inadequate oxygenation in the lungs [35, 36].
6. **Lymphopenia:** Viral infection can lead to marked lymphopenia that can affect both CD4+ and CD8+ T cells [3, 28, 36]. Lymphopenia, reflected by significantly reduced CD4 and CD8 T cells in peripheral blood, is likely due to sequestration and cell death and reflected by significantly reduced CD4 and CD8 T cells in peripheral blood, has been reported in moderate and severe COVID-19 patients. In addition, antigen specific CD8 Cytotoxic T lymphocyte (CTL) responses have been detected approximately a week following viral infection, and the magnitude of the response was observed to have protective or damaging effects [37].
7. **Neutralizing antibodies**: Neutralizing antibodies bind to specific surface receptors on infectious agents such as viruses and toxins, reducing or eliminating their ability to exert harmful effects on cells. SARS-CoV-2 infected individuals generate a robust and long-lasting neutralizing antibody response, and plasma from convalescent COVID-19 patients has been used for treatment of severe disease with some success [38, 39]. It has recently been reported that neutralizing antibodies to SARS-CoV-2 can predict severity and survival, with higher titers being associated with severe disease in some instances [40].

## RESULTS

We evaluated multiple approaches to develop mathematical models using parameters that can predict the progression of disease. Candidate parameters were selected from mechanistic understanding of the process of pathogenesis of COVID-19 to evaluate their possible impact on the clinical outcome. Regression models utilize data to build predictive models. Hypotheses are examined and confirmed with pre-determined statistical confidence and inferential power. These models incorporate all the experimental variability in the data set. Since the models contained numeric factors and numeric ordinal outcomes, we utilized methods of Multiple Linear Regression [41]. In this approach, we used the simulated data set from COVID-19 affected subjects, organized, and analyzed it to understand the variability of each of the parameters.

### Regression modeling approach

The data set was parsed into training and testing partitions using methods of randomization. The validity of the model was based on goodness-of-fit of R Sq. > 90% and ANOVA, [p value <0.05] and a consequent F Ratio **(Table 2a and 2b)**. These statistical results confirmed acceptable degree of predictability of the model.

**Table 2A.**
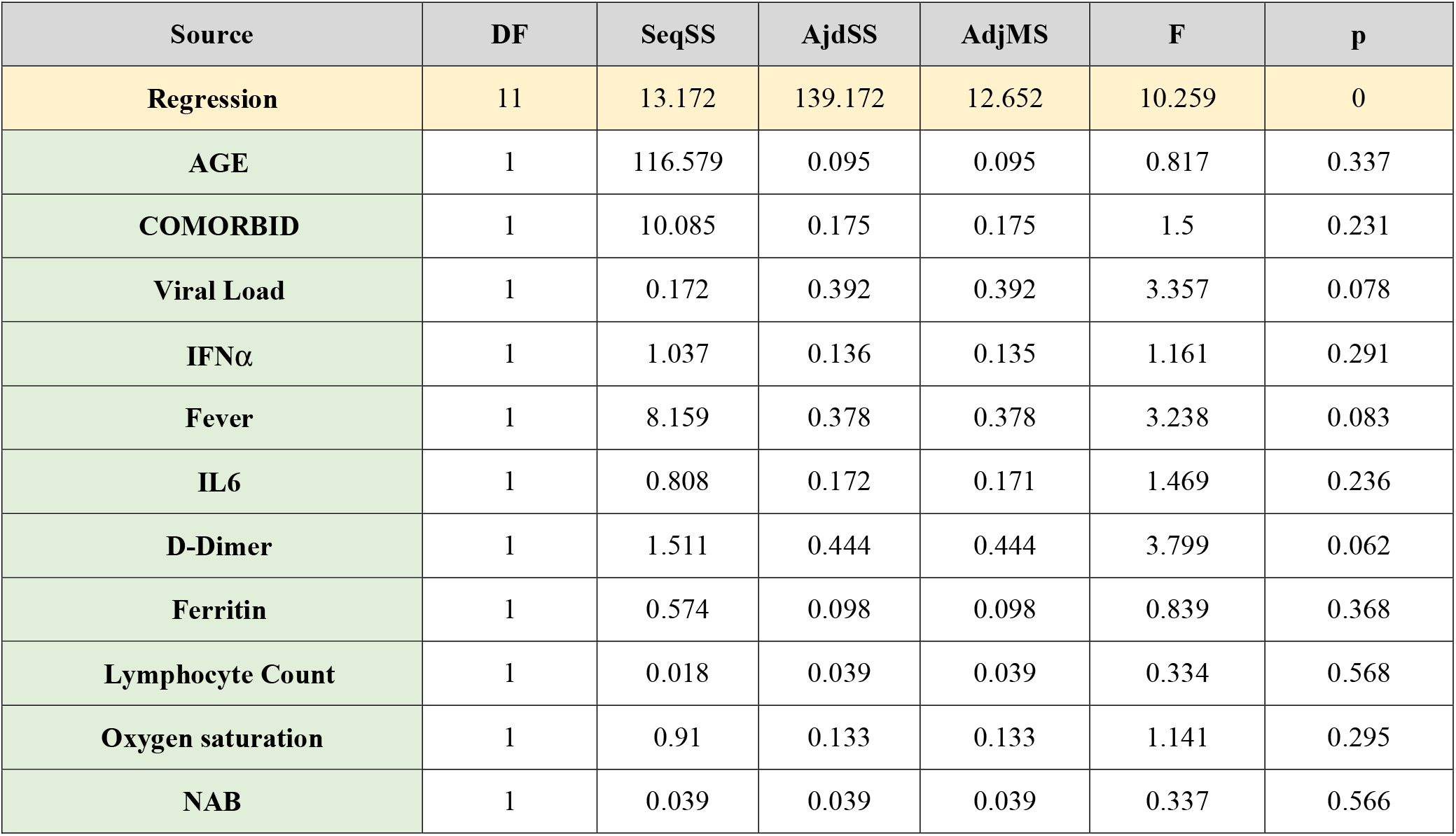
Statistical analysis of coefficients for each parameter based on the multiple regression analysis. The Table shows the regression statistics. The statistical terms are: DF (degrees of freedom); SeqSS (Sequential Sum Square); AdjSS (Adjusted Sum Square), AdjMS (Adjusted Mean Squares), F ratio, p value.

**Table 2B.** Coefficient and standard error for parameters. The coefficients for each parameter were determined by using multiple regression analyses, which is the multiplier to the parameter value in a linear regression equation. The values of the coefficients of each of the parameters are shown using ANOVA. The table shows the standard error, t and p values. The p value denotes statistical significance to the outcome.

| Term | Coefficient | Standard Error | t | p |
| --- | --- | --- | --- | --- |
| Constant | -36.898 | 24.867 | -1.484 | 0.150 |
| AGE | -0.021 | 0.023 | -0.904 | 0.374 |
| COMORBID | 0.894 | 0.730 | 1.225 | 0.232 |
| Viral Load | -0.048 | 0.026 | -1.832 | 0.078 |
| IFN A | -0.005 | 0.005 | -1.077 | 0.291 |
| Fever | 0.444 | 0.247 | 1.799 | 0.084 |
| IL6 | -0.003 | 0.003 | -1.212 | 0.236 |
| D-Dimer | 0.271 | 0.139 | 1.949 | 0.062 |
| Ferritin | 0.000 | 0.001 | 0.916 | 0.368 |
| Lymphocyte Count | -0.001 | 0.001 | -0.578 | 0.568 |
| Oxygen saturation | -0.038 | 0.036 | -1.068 | 0.295 |
| NAB | 0.000 | 0.000 | -0.580 | 0.567 |
| Error | 26.000 | 3.039 | 3.039 | 0.117 |
| Total | 37.000 | 142.211 |  |  |

**Table 3.**
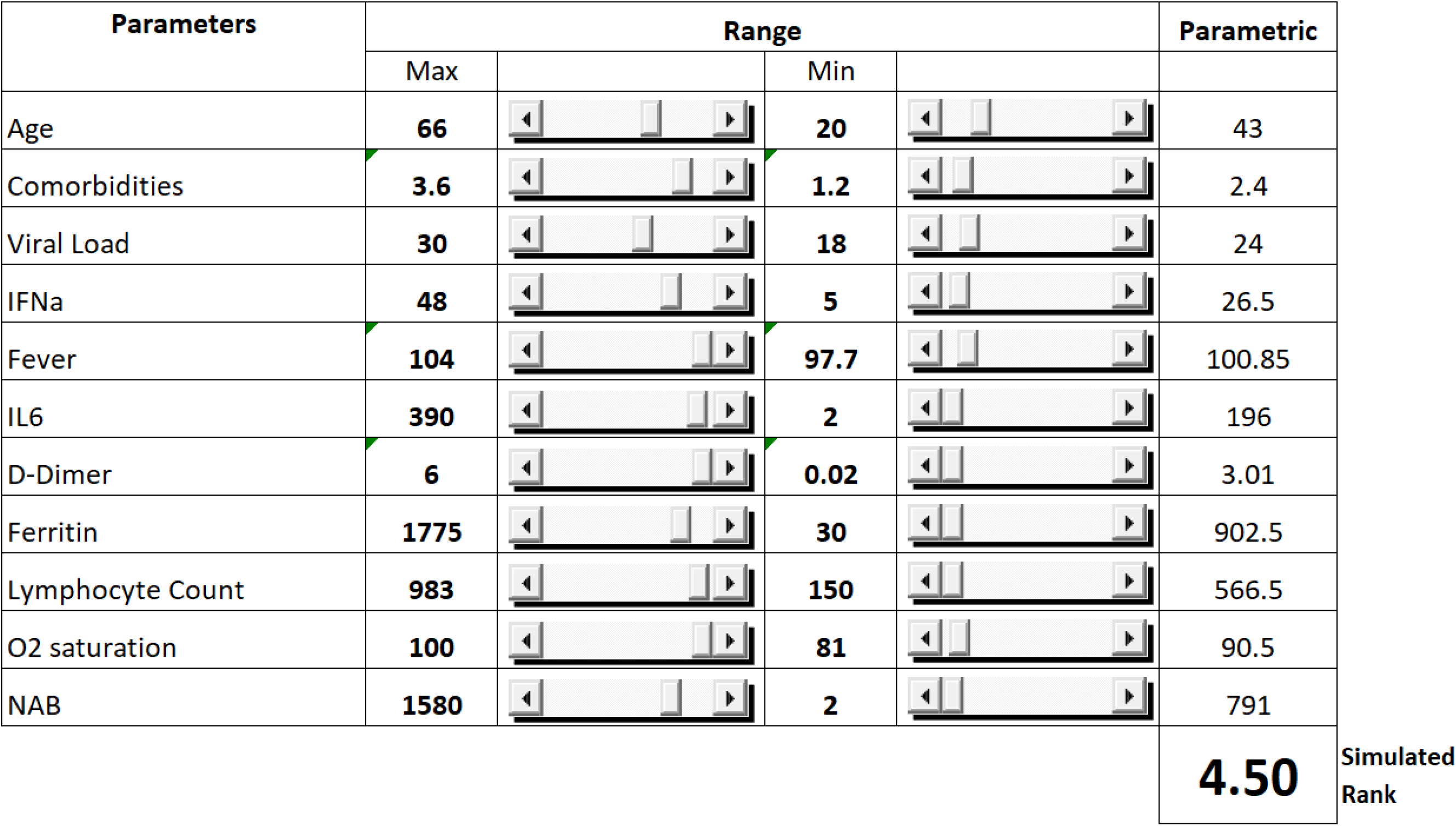
The Ranges for the Monte Carlo Simulation.

| Parameters | Range |  |  |  | Parametric |
| --- | --- | --- | --- | --- | --- |
|  | Max |  | Min |  |  |
| Age | **66** |  | **20** |  | 43 |
| Comorbidities | **3.6** |  | **1.2** |  | 2.4 |
| Viral Load | **30** |  | **18** |  | 24 |
| IFNa | **48** |  | **5** |  | 26.5 |
| Fever | **104** |  | **97.7** |  | 100.85 |
| IL6 | **390** |  | **2** |  | 196 |
| D-Dimer | **6** |  | **0.02** |  | 3.01 |
| Ferritin | **1775** |  | **30** |  | 902.5 |
| Lymphocyte Count | **983** |  | **150** |  | 566.5 |
| O2 saturation | **100** |  | **81** |  | 90.5 |
| NAB | **1580** |  | **2** |  | 791 |
|  |  |  |  |  | **4.50** |
|  |  |  |  |  | Simulated Rank |

Following this multiple-regression analysis, we conducted 2,000 bootstrap samplings using the predicted coefficients and random variates from chosen intervals of parameters. The assumption for this analysis was that each of the parameters were independent variables. The coefficients of each parameter were determined by using multiple regression analyses, which is the multiplier to the parameter value in a linear regression equation. The inclusion of all the variables in analysis ensures their contribution to the model [41]. However, analysts applying this model in the future may, at their judgment, evaluate statistical significance of regression coefficients. Parameters that are not significant maybe excluded using step wise regression. In our analysis, results based on training dataset predictors matched with those from the test dataset confirming an acceptable degree of predictability of the model. We invite the readers of this article to contact us to analyze the predictive potential of the model using their data.

### Monte Carlo simulation

To determine the factors that contribute to the clinical outcome at the population level, Monte Carlo simulation was performed on a sample set of laboratory and clinical parameters covering the full range, from asymptomatic to severe disease, of outcomes (stages 1-7 [Figure 2]) [12, 13]. The histogram and cumulative data (**Figure 3a)** show the distribution of asymptomatic to severe outcomes. The Tornado chart (**Figure 3b**) shows the sensitivity of parameter to the outcome in the selected range.

**Figure 3a.**
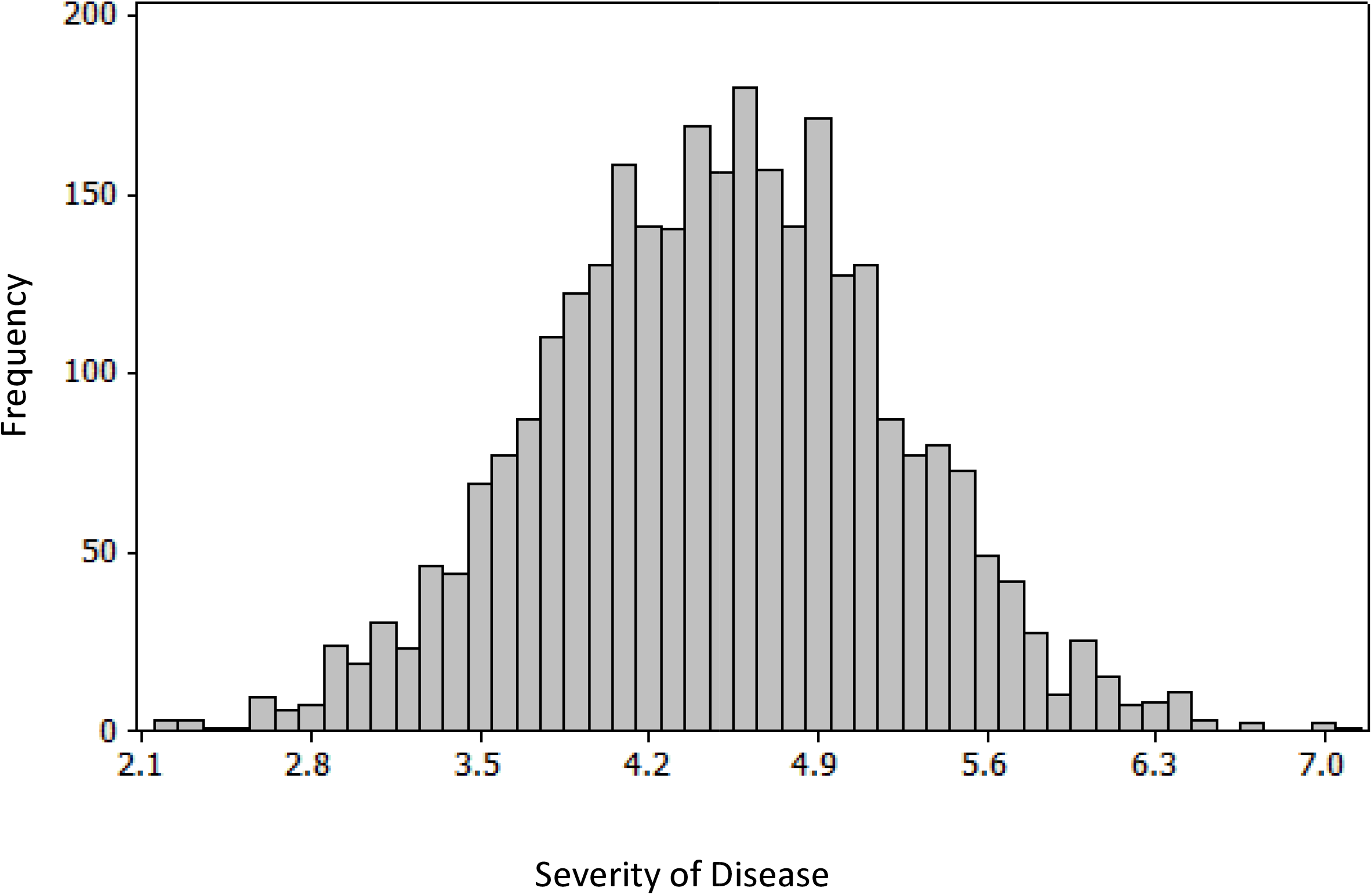
Histogram from Monte Carlo Simulation. 2,000 bootstrap samplings were generated using the predicted coefficients from the linear regression analysis, from the intervals of parameters. The minimum and maximum values for each of the parameters was set to the levels. The distribution of the severity of outcome is in this frequency histogram. The values on the x axis denote the disease severity, and y axis denote frequency of the population in each level of clinical outcome.

**Figure 3b.**
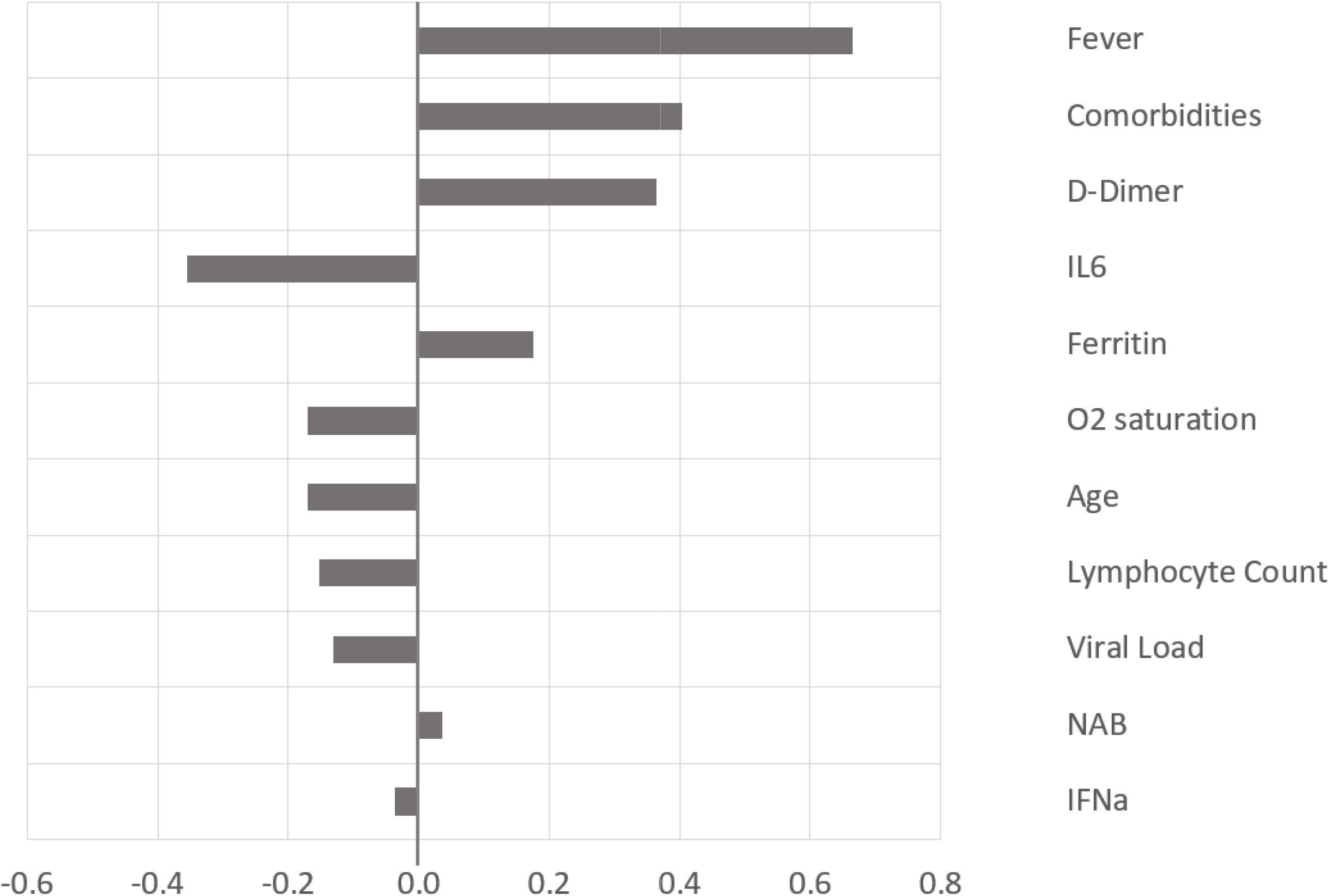
The Tornado Chart. The tornado chart shows the influence of each of the parameters on the outcome. The positive values correlate positively towards the severity of disease, and negative values towards asymptomatic disease.

### The predictive model

Based on the correlation coefficient of the parameters and the outcome from the training data set, we developed a model using the prediction equation. **Table 4** shows the process of predicting the outcome. When the numerical values of the individual parameters for each patient are entered into the columns, the model predicts the outcome. The validation of the model will require data from patients and clinical trials. The goal of this exercise was to develop a model that can be used to predict the outcome in a large number of patients.

**Table 4.** The Prediction of outcome based on observed and predicted values. The values of the parameters for each of the seven subjects are entered in columns, upon running of the model. The predicted values are calculated in numerical values in a range of 1-7, with 1 being asymptomatic, and 7 most severe.

| Variable | Subject 1 | Subject 2 | Subject 3 | Subject 4 | Subject 5 | Subject 6 | Subject 7 |
| --- | --- | --- | --- | --- | --- | --- | --- |
| Age | 20 | 52 | 55 | 55 | 62 | 73 | 80 |
| Comorbidity | 1.1 | 2.2 | 2.3 | 2.4 | 2.6 | 3.2 | 3.5 |
| Viral Load | 36 | 19 | 16 | 15 | 17 | 21 | 17 |
| IFNa | 7 | 20 | 50 | 60 | 40 | 9 | 5 |
| Fever | 98.6 | 99.1 | 99.8 | 100 | 99.4 | 100.1 | 100 |
| IL6 | 1 | 30 | 60 | 70 | 50 | 50 | 90 |
| D-Dimer | 0.05 | 0.25 | 0.40 | 0.45 | 0.35 | 1.5 | 4.0 |
| Ferritin | 30 | 350 | 800 | 1275 | 500 | 1500 | 1800 |
| Lymphocyte Count | 800 | 740 | 620 | 580 | 640 | 340 | 200 |
| Oxygen Saturation | 100 | 95 | 83 | 85 | 90 | 95 | 94 |
| NAB | 2 | 20 | 150 | 200 | 100 | 220 | 400 |
| Calculated Outcome Rank | 0.723 | 3.299 | 4.008 | 4.237 | 3.228 | 4.916 | 6.427 |
| PREDICTED | 1 | 3 | 4 | 4 | 3 | 5 | 6 |
| OBSERVED | 1 | 3 | 4 | 4 | 4 | 5 | 6 |

## DISCUSSION

We have evaluated multiple regression analysis for mathematically modeling the course of COVID-19 to predict clinical outcome. The premise of this model is that quantitatively measured clinical and laboratory parameters involved in the pathogenesis of disease progression can be mathematically mapped to a multiple-regression model. COVID-19 is initiated by infection of the subject with SARS-CoV-2 with subsequent replication in the epithelial cells of the lung. The factors that contribute to the viral load include number of cells that express the ACE2 and other receptors, and inflammatory cytokines. Comorbidities contribute towards a more serious disease progression. Virus infection of antigen presenting cells, such as dendritic cells, macrophages, and other cell types including endothelial cells, results in activation of biochemical signals, which lead to secretion of a battery of cytokines that include IL1β and IL-6. The viral infection as well as inflammatory cytokines cause fever and an increase in serum inflammatory factors such as D-Dimer and Ferritin. Induction of an inflammatory response contributes to reduction of the total numbers of lymphocytes from circulation. The inflammation results in a loss of lung function (e.g., reduction in blood-oxygen levels), cardiac function (blood pressure) and can culminate in multi-organ failure.

Subjects with a normal immune response can generally mount an adequate innate and adaptive response to the virus. These individuals clear the virus by generating adaptive T cell responses and neutralizing antibodies. Subjects with comorbid conditions can have compromised immune function which could result in dysfunctional activation of inflammatory responses, leading to worse clinical outcomes.

Selection of the parameters that were included in the model building process was influenced by their perceived significance from current research reports. This list of factors is by no means complete and it is expected that in due course a more comprehensive list will emerge. This report provides a basis for creating a tool, independent of the number and type of parameters, that could find utility in predicting the disease outcome using those parameters.

### Viral Load

Association of viral load and progression of diseases has been reported for several viral infections [42-44]. Viral load in COVID-19 is measured by qRT-PCR of SARS-CoV-2 using primers for the spike gene [43]. The correlation of high viral load with severity of disease progression has been extensively demonstrated. The systemic dissemination of the virus has been associated with expression of the ACE2 receptor on endothelial cells [21]]. Comorbid conditions could enhance the expression of receptors and enable distribution of virus, thereby enhancing the viral load, which can result in progression of disease.

### IFNα

The critical role of Type I interferons in innate and adaptive immunity, leading to both protective and pathogenic responses, has been reported in the case of several viral and bacterial infections [45]. SARS-CoV-2 infection has been shown to result in a diverse range of effects on Type I immune responses. Most patients elicit a strong IFNα response along with a battery of inflammatory cytokines, some of which progress to a cytokine storm [46, 47]. Specific blocking of the type I mediated signal transduction by various proteins of SARS-CoV-2 has been demonstrated [48]. A remarkably high proportion of male subjects experiencing severe or critical COVID-19 disease expressed an inability to produce sufficient levels of IFNα due to various types of errors in the IFN genes. Curiously, majority of the male subjects possessed circulating IFNα autoantibodies that had the ability to neutralize the endogenously produced cytokine, thereby effectively reducing the available IFNα. The discovery of these two mechanisms for lowering IFNα levels underscores its relevance in controlling the progression of disease in individuals infected with the SARS-CoV-2 [49].

### D-Dimer

D-Dimer is routinely measured in clinical situations because its levels correlate with serious underlying conditions including venous thromboembolism, cancer and sepsis [48]. In the case of COVID-19 patients, introduction of the virus brings about infection-induced inflammatory alterations leading to coagulopathy. Lungs being the target of SARS-CoV-2, acute injury to the lung as well as multi organ failure have been caused by the virus-induced cascade of the inflammatory pathway. In an early study on 41 COVID-19 patients, those with severe disease had higher levels of D-Dimer along with high levels of IL-8, TNFα and IL-2R [31]. Male patients were found to have higher levels of IL-6, IL-2R, Ferritin and other markers of inflammation compared to female. High levels of IL-6 showed a statistically significant correlation with severe disease in a retrospective study as well [27]. One can hypothesize that such patients would likely benefit from anticoagulation therapy.

### Ferritin

A high level of ferritin, measure of stored iron, was found to be associated with severe disease in COVID-19 patients and was linked to high fatality rates in a 72 patient prospective study [33, 50, 51]. In another study on 39 patients, those with mild COVID-19 symptoms had lower levels of ferritin while those with moderate or severe symptoms expressed higher levels of ferritin [50].

### Lymphopenia

Loss of lymphocytes after viral infections has been associated with severe disease. The mechanisms involved in lymphodepletion can been implicated to be due to cell death, cytokine storm and/or redistribution of lymphocyte populations [3, 33, 37]. In this model, we have utilized lymphopenia as a measure of severity of disease progression. Loss of immune function could result in several potential mechanisms of pathogenesis including autoimmunity, hyperactivation, increased susceptibility to infections and organ dysfunction.

### Neutralizing Antibodies

Induction of neutralizing antibodies directed to the receptor-binding domain of the spike protein is critical for restricting entry of the virus into the cells and has been one of the central tenets of a protective immune response. In this model, we have used a range of IgG titers to spike protein for the simulated data set [52]. However, the role of neutralizing antibodies induced in a large proportion of subjects following natural infection is still being studied [53]. Some subjects do not elicit strong antibody responses. Sub-optimal levels of antibodies may catalyze generation of virus mutants [54]. Neutralizing antibodies to the virus have generally not correlated with reduced severity of disease in the primary infection. In addition, it will be interesting to decipher the role of pre-existing antibodies reported recently in the modulation of disease and its impact on vaccination regimens. Thus, the mechanisms involved in the induction of antibodies, the repertoire and diversity of responses, and effects on protection versus progression, remains to be clearly established.

The predictive model can have multiple applications, such as forecasting the percentage of the population that will progress to severe disease in each geography, enabling logistics planning for hospital beds, health care providers and personal-protective safety equipment. Analysis of the coefficient of correlations of parameters with outcome of disease may provide clues to a better understanding of the mechanism of action of disease pathogenesis. The model can predict the probability of disease progression at an individual level, based on parameter data, and can be used to understand the effect and impact of therapeutic interventions. The predictive model can be utilized to analyze large amounts of data to develop algorithms for personalized treatment regimens.

In summary, we have developed a probabilistic model that can be utilized to predict progression of disease following infection with SARS-CoV-2. This model was developed using simulated data based on published levels of COVID-19 related clinical and laboratory parameters and provides an approach to predicting the outcome of disease. Validation of the model will require existing data and the clinical outcomes of patients. Prediction of disease progression can be highly valuable at an individual as well as population level.

## Data Availability

The data in the paper will be available.

## ACKNOWLEDGEMENTS

VRN is partly supported by a grant (P30GM114737) from the Centers of Biomedical Research Excellence, National Institute of General Medicine, National Institutes of Health.

